# Harnessing the 100,000 Genomes Project whole genome sequencing data - an unbiased systematic tool to filter by biologically validated regions of functionality

**DOI:** 10.1101/2020.03.30.20047209

**Authors:** Sihao Xiao, Zhentian Kai, David Brown, Genomics England Research Consortium, Claire L Shovlin

## Abstract

Whole genome sequencing (WGS) is championed by the UK National Health Service (NHS) to identify genetic variants that cause particular diseases. The full potential of WGS has yet to be realised as early data analytic steps prioritise protein-coding genes, and effectively ignore the less well annotated non-coding genome which is rich in transcribed and critical regulatory regions. To address, we developed a filter, which we call GROFFFY, and validated in WGS data from hereditary haemorrhagic telangiectasia patients within the 100,000 Genomes Project. Before filter application, the mean number of DNA variants compared to human reference sequence GRCh38 was 4,867,167 (range 4,786,039-5,070,340), and one-third lay within intergenic areas. GROFFFY removed a mean of 2,812,015 variants per DNA. In combination with allele frequency and other filters, GROFFFY enabled a 99.56% reduction in variant number. The proportion of intergenic variants was maintained, and no pathogenic variants in disease genes were lost. We conclude that the filter applied to NHS diagnostic samples in the 100,000 Genomes pipeline offers an efficient method to prioritise intergenic, intronic and coding gDNA variants. Reducing the overwhelming number of variants while retaining functional genome variation of importance to patients, enhances the near-term value of WGS in clinical diagnostics.

## INTRODUCTION

Whole genome sequencing (WGS) has become part of the medical genetic repertoire in the UK, but currently, the majority of its potential is unrealised. DNA-derived signals integrate to determine not only the disease phenotypes of specific interest, but also an organism’s growth, development, and responses to environmental challenges including therapeutics. In any one individual, WGS identifies millions of DNA variants compared reference sequences. These are present in just under 20,000 protein-coding genes, and also in much less understood regions of the genome that have diverse functions including transcription into noncoding RNAs, participation in DNA chemical changes that modify transcription, and binding to other nucleic acids or proteins.[1,2]

Current WGS clinical foci are almost exclusively on a subgroup of protein-coding genes where biological function is already known. DNA variants in disease-causing genes are classified by structured methods that assign variant alleles on a spectrum from benign/likely benign, through variant of uncertain significance (VUS) to likely pathogenic/pathogenic for any relevant disease.[3] In research spheres, in order to reduce the number of variables per sample, interrogation of WGS data also usually commences with prioritization methods based on selection of genes, exons, specific genomic regions, patterns of inheritance or phenotypic associations. Variants in the non-coding genome, while not pre-depleted by the sequencing methodology, are effectively deleted in the early analytic stages of variant prioritisation.

This is understandable when the challenges are considered. While the noncoding genome is rich in transcribed and critical regulatory regions, in multicellular eukaryotic organisms such as man, for a similar number of protein-coding genes compared to prokaryotes, there is a vastly expanded genome size. Duplicated, repetitive and other new genomic DNA sequences are biologically prone to variation.[4] Furthermore, the relatively short next-generation sequencing reads mean such regions are also sources of artefactual variation arising from multiple potential alignments of WGS outputs.[5]

We hypothesised that it would be possible to use design a more efficient variant prioritisation method for WGS and apply to NHS-based WGS sequencing outputs. Markers of epigenetics and DNA-protein interactions have been applied genome-wide by molecular laboratories, and an enormous body of biological experimental data made publicly available. As a result, there now exist repositories of information indicating which sections of DNA are more or less likely to have a functional role in at least one examined tissue.[6-10] Similar repositories define DNA loci where repetitive regions impede unique alignments to specific genomic sequences.[5,11] Thus although there is no accurate map of all functional genomic regions in human genomes, and it is difficult to predict *a priori*, where all regulatory elements for a specific gene locus would be located, we hypothesised that it would be possible to use the available datasets to make better use of complete WGS outputs.

Here we report the design and validation of a method that we have applied within the 100,000 Genomes Project, to prioritise non coding variants with more confidence of validity and function, while retaining pathogenic variants identified in coding genes.

## METHODS

### Defining test and validation datasets through the 100,000 Genomes Project

The 100,000 Genomes Project was set up by the UK Department of Health and Social Security in 2013, to sequence whole genomes from National Health Service (NHS) patients recruited through one of 13 Genomic Medicine Centres (GMCs). For successfully nominated Rare Diseases and Cancers, patient/family DNA sampling was accompanied by phenotypic data entries and integration with NHS health records. Anonymised raw sequencing data were made available in the Genomics England Research Environment.[12] Separately, Genomics England performed data alignments, and variant classifications for the clinical diagnostic pipeline: For known disease-causing genes, loss of function variants were defined as Tier 1, other variants as Tier 2 variants. Variants in other protein coding genes not in the virtual gene panel for the disease or any listed patient phenotype were classified as Tier 3 variants.

In order to provide test and validation sets, we focused on WGS data from patients with one particular rare disease, hereditary hemorrhagic telangiectasia (HHT).[13] Due to specific circumstances in NHS commissioning, a group of HHT patients who had not previously undergone genetic testing had been recruited, and a subset of these provided a validation dataset, as the Tiering process identified variants in known disease-causing genes.

### Generating GROFFFY

Genomic coordinates of regions included in the GROFFFY were generated from publicly available databases using the Imperial College High Performing Computing service. As our goal was to miss no functional information, wherever possible, experimentally-derived biological data were used in preference to computational predicted files with potential for false negatives. Genomic coordinates were extracted from data aligned to GRCh38 genome build[14], and excluded data originating in cancer cells. Full details are provide in the online Data Supplement (*Supplementary Methods and Tables 1,2*). Briefly:

> ***Genomic coordinates for transcribed loci*** including protein-coding genes, pseudogenes and long non-coding RNAs (lncRNA) were extracted from GENCODE Version 31[10]. LncRNA coordinates were also downloaded from lncipedia version 5.2,[15] and micro RNAs (miRNA) coordinates downloaded from miRbase release 22.1.[16]
>
> ***Genomic coordinates for candidate regulatory element (cRE) regions*** were generated by merging 18,828 bed files identifying regions where DNA had interaction with different elements, such as histone modifications, protein binding, and nuclease accessibility. Thus, DNA binding regions from 2,823 experiments performed in 9 different labs, 4,409 bed files identifying representative DNase hypersensitivity sites from 631 experiments, contained within the ENCODE Encyclopedia registry [7] were downloaded, together with coordinates for CpG islands in the UCSC database.[9]
>
> ***Genomic coordinates of regions excluded*** were based on the ENCODE blacklist (accession ENCSR636HFF) [5] downloaded from ENCODE,[6,7] and RepeatMasker downloaded from the UCSC database [8,9].

### Analysis of Whole Genome Sequencing Data

The final genomics coordinates for GROFFFY were transferred into the Genomics England Research Environment. The full pipeline employed for the current manuscript is illustrated as *Supplementary Figure 1*. At all stages, the number of target set DNA variants after exclusion was recorded and outputted to log files. Stepwise, filter by value functions were written to apply allele frequency filters (excluding variants where general allele frequency exceeded 0.0002[3] in either the 1000 Genome Project[17] or gnomAD database[18]); to exclude synonymous variants (note that synonymous variants within splice regions were retained as they were annotated as “splice region” variants); and to remove additional variants in annotated WGS files for the 34 “Validation Set” HHT DNAs where a causative variant had already been identified in known HHT genes through Tiering. Combined Annotation-Dependent depletion score annotation, using version 1.5 of the CADD database,[19] was quite slow, and the process was put towards the end of the analysis pipeline, so that there were fewer variants that needed to be annotated.

**Figure 1:**
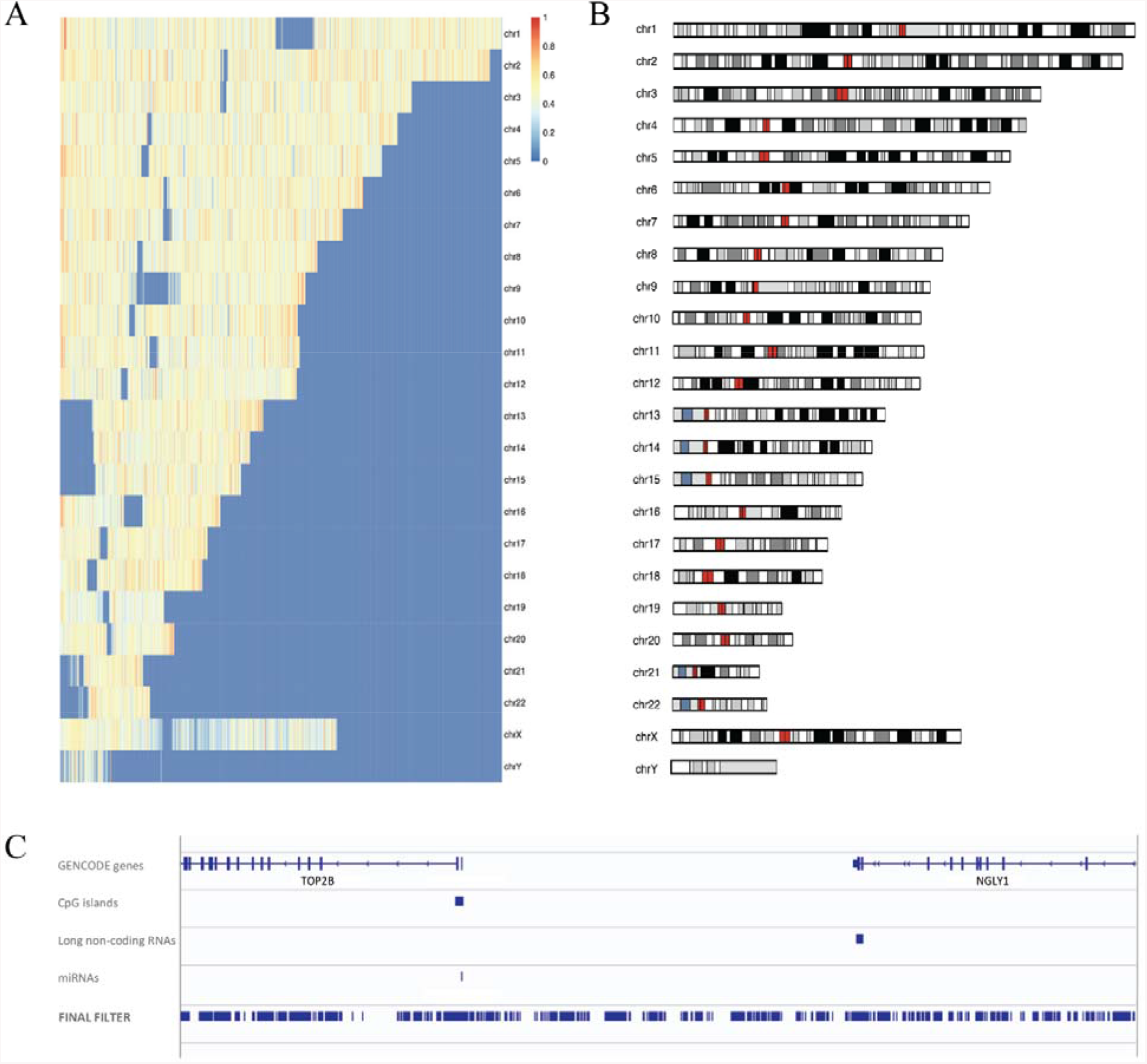
GROFFFY categorization of the human genome: **A)** Heatmap displaying 61,799 data points at 500kb resolution mapped against GROFFFY. Colors represent the percentage of each 500kb region included in GROFFFY with red highest (> 90%), orange ∼70%, yellow ∼40%, and blue <30%. All chromosomes are left-aligned, and as all are shorter than chromosome 1, the right-aligned blue regions indicate the absence of chromosomal material for the indicated chromosome. Other broad blue regions highlight regions particularly devoid of genes and regulatory elements, for example the pericentromeric long arms of chromosomes 1 and 9, and the short arms of chromosomes 13, 14, 15. **B)** The human karyotype for comparison, plotted in R using sourced ideograms with centromeres highlighted in red. (Note consistency with sizes in A). **C)** Randomly chosen region of chromosome 3 (chr3:25,597,986-2-25,783,443). The top 4 tracks illustrate GENCODE gene annotations, CpG islands, long non-coding RNAs, and miRNAs. The final filter is shown as the lowest track. Note this contains both intra and intergenic regions for the region, and that the raw data were not subjected to any processed annotation tracks.

## RESULTS

### GROFFFY defines biologically validated regions of functionality in the human genome

GROFFFY essentially excludes biologically less important regions of the genome by only including regions where biological experiments have generated evidence in favour of functional roles. Due to many overlapping regions, merged bed files were approximately 10-fold the size of the human genome (e.g. for DNA-binding regions, 49,941,695 Kb) before sorting. Following sorting, the GROFFFY filter region based on positive selection of transcribed loci and candidate regulatory element (cREs), and masking of repetitive regions, included 44.4% of the human genome. Genome-wide illustration of GROFFFY by heat mapping datapoints at 500kb resolution highlighted genomic regions that are enriched or devoid of genes and regulatory elements (*Figure 1A, 1B*). To illustrate GROFFFY at higher resolution, we randomly selected a 185kb region containing two protein-coding genes. *Figure 1C* illustrates the raw experimental data of the GROFFFY region as the lowest track.

### GROFFFY prioritisation of variants within WGS data

GROFFFY was applied to WGS-derived DNA variants in individuals with HHT [13] recruited to the 100,000 Genomes Project. The experimental target dataset comprised 98 whole genomes. The validation dataset included 34 whole genomes where a disease-causing variant had already been identified through clinical diagnostic pipelines.

We focused first on the experimental target dataset of 98 whole genomes. The scale of the bioinformatics challenge was emphasized by a total of 26,271,838 unique changes compared to the human genome reference sequence (GRCh38 genome build [14]). Pre filtration, the number of DNA variants per individual target dataset DNA ranged from 4,786,039 to 5,070,340 (mean 4,867,167, SD 48,868), and approximately one third (32.28%), lay within intergenic areas.

Applying GROFFFY as a first filter reduced the mean number of variants by 2,812,015 per DNA, while retaining variants within intergenic areas (*Supplementary Table 3A, Supplementary Figure 2A*). Post filtration, intergenic variants represented 33.02% of the total variant number.

**Figure 2:**
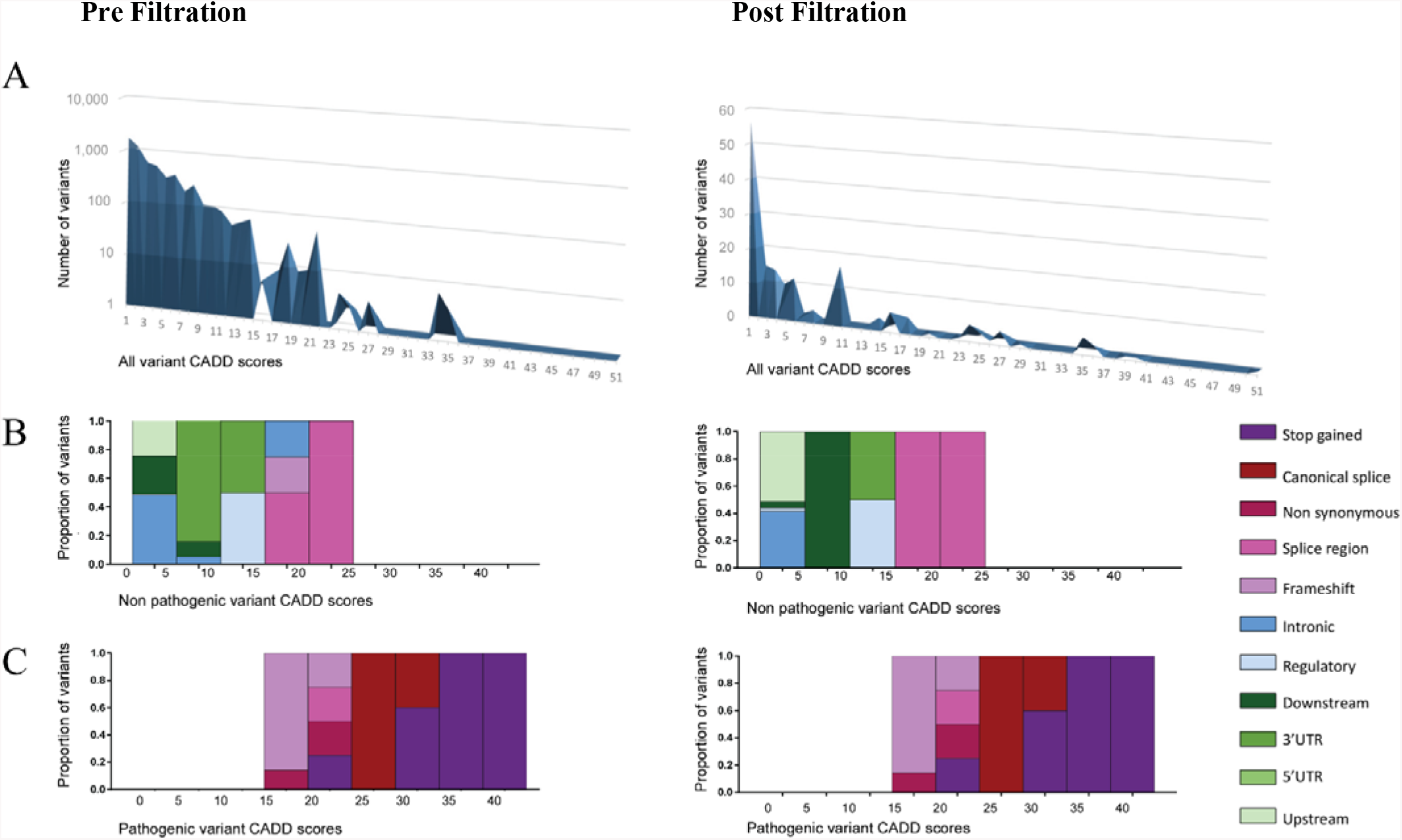
GROFFFY validation by retention of pathogenic variants: Comparison of variants in the 34 validation dataset whole genomes by Combined Annotation-Dependent depletion (CADD) scores indicative of likely pathogenicity pre, and post filtration. **A)**: Total number of variants with indicated CADD scores (note logarithmic scale pre filtration versus linear scale post filtration. **B):** Non-pathogenic variants by CADD score categories, and molecular subtype as noted in key. **C)**: Pathogenic variant scores by CADD score categories, and molecular subtype as noted in key. Note plots in C are identical, as all variants were still present post filtration.

We next tested as if seeking a single rare, disease-causing, pathogenic variant in each target dataset DNA. In keeping with recommendations for rare disease pathogenicity assignments,[3] 2×10^−4^ was set as the population allele frequency threshold to be included: Referencing allele frequencies in the 1000 Genome Project[17] and gnomAD[18] removed a mean of 2,476,589 and 2,483,377 variants per DNA respectively (*Supplementary Figure 2A*). Further rare-variant specific filters were then employed to exclude (i) the non-pathogenic variants present in the validation dataset DNAs (“white listed variants”), (ii) synonymous variants that were not in splice or regulatory regions, and (iii) variants where deleteriousness was not predicted based on low Combined Annotation-Dependent depletion (CADD) scores.[19] These filters removed a mean of 307,152 further variants. Overall, following application of all filters, a mean of 21,486 (0.44%) of variants were left per DNA, totaling 873,554 different variants across all DNAs. Comparing the pre and post filter variant numbers generated a Wilcoxon rank-sum p-value of <2.2e^-16^. Irrespective of other filters applied, GROFFFY, and its individual components, significantly reduced the number of variants compared to the other tested filter sets (*Supplementary Table 3B, Supplementary Figure 2B*).

### Validating GROFFFY

Finally, we demonstrated that applying GROFFFY did not delete key variants by examining the validation dataset. These 34 DNAs each harbored a single variant in one of the known genes for HHT, a vascular dyplasia caused by a single null allele most commonly in *ENG, ACVRL1* or *SMAD4*.[13] With the exception of a mosaic case identified only by the clinical diagnostic pipelines (and hence not present in the unfiltered dataset[20]), all pathogenic variants were retained post filtration (*Figure 2*). identicality of plots as all variants were still present post filtration.

## DISCUSSION

In this manuscript we present and validate a simple code that synthesises biologically-generated signals of function in order to filter out variants in DNA regions with no such evidence of functionality. This generically applicable method was highly effective in reducing the number of variants from WGS performed within the 100,000 Genomes Project from almost 5 million per individual to an average of about 21,000. GROFFFY retained intergenic, intronic and coding gDNA variants, and critically, retained key pathogenic variants already known from a validation dataset. The approach is thus expected to enhance discovery by enabling further analyses to focus on a smaller number of variants more likely to be implicated in generating a phenotype.

Other variant filtration methods are already used in WGS, with many depending on union and intersection rules of existing annotation tracks from ENCODE, ENCODE Encyclopedia and UCSC databases. The candidate cis-regulatory elements file produced by ENCODE is particular favored with its specific predictions of each possible CRE position and size. By using the raw biological data providing broader areas for inclusion, GROFFFY is proposed to better suit the purpose of a first pass filter for definition of variants worthy of further study, than computational predicted files with potentially high false negative rates.

For researchers wishing to apply open access WGS data to their own areas of research, GROFFFY stringency could be further increased according to a researcher’s tissue of interest by restricting the “selected for” regions only to the specific tissue type. Nevertheless, even the least stringent filter efficiently restricts the number of variants identified by WGS towards a number more suited to machine and other computational approaches

In conclusion, we present and validate a filter that reduces the overwhelming number of variants identified by WGS, while retaining functional genome variation of importance to patients. Retention of non-coding variants enhances the near-term value of WGS and supports wider use of WGS in clinical diagnostics.

## Data Availability

Primary data in an anonymised format are available to Researchers within the Genomics England Research Environment.

https://www.genomicsengland.co.uk/about-gecip/joining-research-community/

## Acknowledgements

This research was made possible through access to the data and findings generated by the 100,000 Genomes Project. The 100,000 Genomes Project is managed by Genomics England Limited (a wholly owned company of the Department of Health and Social Care). The 100,000 Genomes Project is funded by the National Institute for Health Research (NIHR) and NHS England. The Wellcome Trust, Cancer Research UK and the Medical Research Council have also funded research infrastructure. The 100,000 Genomes Project uses data provided by patients and collected by the National Health Service as part of their care and support. We thank the National Health Service staff of the UK Genomic Medicine Centres and the participants for their willing participation. This research was co-funded by the NIHR Imperial Biomedical Research Centre. The views expressed are those of the authors and not necessarily those of funders, the NHS, the NIHR, or the Department of Health and Social Care.

## Contributorship Statement

SX devised and generated the GROFFFY approach, devised all scripts to generate GROFFFY, and generated all numeric data, Figure 1 and Supplementary Tables and Figures. ZK advised on Linux and script generation. DB set up and advised on the Research Environment. CLS devised concepts, advised on approaches and data analysis, generated Figure 2, and wrote the manuscript. All authors reviewed and approved the final manuscript.

## REFERENCES

1. Ransohoff JD, Wei Y, Khavari PA. 2018. The functions and unique features of long intergenic non-coding RNA. Nat Rev Mol Cell Biol 19(3):143–157.

2. Marchal C, Sima J, Gilbert DM. 2019. Control of DNA replication timing in the 3D genome. Nat Rev Mol Cell Biol 20(12):721–737.

3. Richards S, Aziz N, Bale S, Bick D, Das S, Gastier-Foster J, Grody WW, Hegde M, Lyon E, Spector E, Voelkerding K, Rehm HL; ACMG Laboratory Quality Assurance Committee. 2015. ACMG Laboratory Quality Assurance Committee. Standards and guidelines for the interpretation of sequence variants: a joint consensus recommendation of the American College of Medical Genetics and Genomics and the Association for Molecular Pathology. Genet Med. 17(5):405–24

4. Janssen A, Colmenares SU, Karpen GH. 2018. Heterochromatin: Guardian of the Genome. Annu Rev Cell Dev Biol 34:265–288.

5. Amemiya HM, Kundaje A, Boyle AP. The ENCODE Blacklist: Identification of Problematic Regions of the Genome. 2019. Sci Rep 9(1):9354

6. Sloan CA, Chan ET, Davidson JM, Malladi VS, Strattan JS, Hitz BC, Gabdank I, Narayanan AK, Ho M, Lee BT, Rowe LD, Dreszer TR, Roe G, Podduturi NR, Tanaka F, Hong EL, Cherry JM. 2016. ENCODE data at the ENCODE portal. Nucleic Acids Res 44(D1):D726–32

7. Davis CA, Hitz BC, Sloan CA, Chan ET, Davidson JM, Gabdank I, Hilton JA, Jain K, Baymuradov UK, Narayanan AK, Onate KC, Graham K, Miyasato SR, Dreszer TR, Strattan JS, Jolanki O, Tanaka FY, Cherry JM. The Encyclopedia of DNA elements (ENCODE): data portal update. 2018. Nucleic Acids Res 46(D1):D794–D801, accessed for data 25.07.2019

8. Kent WJ, Sugnet CW, Furey TS, Roskin KM, Pringle TH, Zahler AM, Haussler D. 2002. The human genome browser at UCSC. Genome Res. 12(6):996–1006.

9. Haeussler M, Zweig AS, Tyner C, Speir ML, Rosenbloom KR, Raney BJ, Lee CM, Lee BT, Hinrichs AS, Gonzalez JN, Gibson D, Diekhans M, Clawson H, Casper J, Barber GP, Haussler D, Kuhn RM, Kent WJ. 2019 The UCSC Genome Browser database: 2019 update. Nucleic Acids Res 47(D1):D853–D858, accessed for data 11.03.2019.

10. Harrow J, Denoeud F, Frankish A, Reymond A, Chen CK, Chrast J, Lagarde J, Gilbert JG, Storey R, Swarbreck D, Rossier C, Ucla C, Hubbard T, Antonarakis SE, Guigo R. 2006. GENCODE: producing a reference annotation for ENCODE. Genome Biol 7 Suppl 1:S4.1-9.

11. Institute for Systems Biology Repeatmasker available at: http://www.repeatmasker.org, accessed for data 25.09.2019

12. Research Environment User Guide - Genomics England Research Environment - Genomics England Confluence. Available at: https://cnfl.extge.co.uk/display/GERE/Research+Environment+User+Guide

13. European Reference Network For Vascular Diseases (VASCERN-HHT): Orphanet 2019 definition of hereditary hemorrhagic telangiectasia. Available at. https://www.orpha.net/consor/cgi-bin/OC_Exp.php?Expert=774, accessed 19/11/2019

14. Genome Reference Consortium Human Build 38. Available at https://www.ncbi.nlm.nih.gov/assembly?term=GRCh38&cmd=DetailsSearch

15. Volders P-J, Anckaert J, Verheggen K, Nuytens J, Martens L, Mestdagh P, Vandesompele J. LNCipedia 5: towards a reference set of human long non-coding RNAs. Nucleic Acids Res 2019;47(D1):D135–D139.

16. Kozomara A, Birgaoanu M, Griffiths-Jones S. miRBase: from microRNA sequences to function. Nucleic Acids Res 2019; 47(D1):D155–62

17. Zheng-Bradley X, Streeter I, Fairley S, Richardson D, Clarke L, Flicek P; 1000 Genomes Project Consortium. 2017. Alignment of 1000 Genomes Project reads to reference assembly GRCh38. Gigascience 6(7):1–8.

18. Lek M, Karczewski KJ, Minikel EV, Samocha KE, Banks E, Fennell T, O’Donnell-Luria AH, Ware JS, Hill AJ, Cummings BB, Tukiainen T, Birnbaum DP, Kosmicki JA, Duncan LE, Estrada K, Zhao F, Zou J, Pierce-Hoffman E, Berghout J, Cooper DN, Deflaux N, DePristo M, Do R, Flannick J, Fromer M, Gauthier L, Goldstein J, Gupta N, Howrigan D, Kiezun A, Kurki MI, Moonshine AL, Natarajan P, Orozco L, Peloso GM, Poplin R, Rivas MA, Ruano-Rubio V, Rose SA, Ruderfer DM, Shakir K, Stenson PD, Stevens C, Thomas BP, Tiao G, Tusie-Luna MT, Weisburd B, Won HH, Yu D, Altshuler DM, Ardissino D, Boehnke M, Danesh J, Donnelly S, Elosua R, Florez JC, Gabriel SB, Getz G, Glatt SJ, Hultman CM, Kathiresan S, Laakso M, McCarroll S, McCarthy MI, McGovern D, McPherson R, Neale BM, Palotie A, Purcell SM, Saleheen D, Scharf JM, Sklar P, Sullivan PF, Tuomilehto J, Tsuang MT, Watkins HC, Wilson JG, Daly MJ, MacArthur DG; Exome Aggregation Consortium. 2016. Analysis of protein-coding genetic variation in 60,706 humans. Nature 536:285–91

19. Rentzsch P, Witten D, Cooper GM, Shendure J, Kircher M. 2019. CADD: predicting the deleteriousness of variants throughout the human genome. Nucleic Acids Res 47(D1):D886–D894.

20. Clarke JM, Alikian M, Xiao S, Kasperaviciute D, Thomas E, Turbin I, Rose G, Olupona K, Cifra E, Curetean E, Ferguson T, Redhead J, Genomics England Research Consortium, Shovlin CL. 2020. Low grade mosaicism in hereditary haemorrhagic telangiectasia identified by bidrectional whole genome sequencing reads through the 100,000 Genomes Project clinical diagnostic pipeline. J Med Genet 2020 in press.

